# Spinal Cord Versus Brain Imaging Biomarkers of Multiple Sclerosis Trajectory Combining 7T and 3T MRI

**DOI:** 10.1101/2025.06.13.25329482

**Authors:** Alessandro Miscioscia, Constantina A. Treaba, Elena Barbuti, Valeria T. Barletta, Jacob Sloane, Eric C Klawiter, Julien Cohen-Adad, Paolo Gallo, Patrizia Pantano, Caterina Mainero

## Abstract

**Background:** In multiple sclerosis (MS), 7 Tesla (7T) MRI improves the visualization of cortical (CLs) and white matter (WM) lesions with a paramagnetic rim (PRLs), associated with smoldering inflammation. Spinal cord (SC) atrophy is a critical determinant of clinical disability in MS, but its importance relative to PRLs and CLs in predicting neurological disability remains unclear.

**Purpose:** To identify the most relevant predictors for baseline neurological disability and 4-year disease progression independent of relapse activity (PIRA) in a heterogeneous MS cohort.

**Materials and Methods:** One-hundred-twelve MS patients (83 relapsing-remitting, 29 secondary progressive) were prospectively recruited between 2010 and 2024. 7T T2*-susceptibility-weighted imaging was acquired to segment CLs, PRLs, and non-rim WM lesions, and 3T T1-weighted brain MRI to estimate cortical thickness, brain WM volume, and the SC C2-C3 cross-sectional area (CSA) using FreeSurfer and Spinal Cord Toolbox. Expanded Disability Status Scale (EDSS) was assessed at baseline and longitudinally, in 97/112 MS patients, after a mean follow-up of 4.0 years. Associations between imaging metrics and clinical outcomes were evaluated using regression models.

**Results:** Baseline EDSS was associated with non-rim WM lesion volume (p=<0.001), CL volume (p=0.001), brain WM volume (p=0.017), and C2-C3 CSA (p=0.003). At follow-up, 23/97 patients showed PIRA. PIRA was associated with PRL volume (p=0.030), CL volume (p=0.011), and brain WM volume (p<0.001). A stepwise logistic regression identified CL volume as the strongest independent predictor of PIRA (Nagelkerke R^2^=0.200, OR=1.0006, p=0.005). Patients with a CL load > 403 mm^3^ progressed in half of cases (70% sensitivity, 50% specificity) within 4 years.

**Conclusion:** In MS, different imaging biomarkers are associated with either the current disability or PIRA. Spinal cord atrophy mainly explains the current EDSS, while brain WM atrophy and PRLs provide additional insights into future disability trajectory. Among all markers, CLs emerged as the main driver for PIRA.

## Introduction

Multiple sclerosis (MS) is a chronic inflammatory, demyelinating, and neurodegenerative disease of the central nervous system (CNS), and the most common cause of neurological disability in young adults after trauma (1). Disability accumulation in MS can result from either incomplete recovery following relapses (relapse-associated worsening [RAW]) or progression that occurs independently of relapse activity (PIRA) (2). MRI biomarkers of brain and spinal cord (SC) damage can provide crucial information about the trajectory of clinical disability in MS (3). Particularly, paramagnetic rim lesions (PRLs), cortical lesions (CLs), as well as volumetric measures of SC were shown to be strong predictors of PIRA (4–8). In a previous study, we identified PRL and CL volumes as top predictors of Expanded Disability Status Scale (EDSS) progression (7), while SC involvement was not assessed.

Although the individual contributions of SC abnormalities and CLs relative to PRLs have been previously investigated (5,6), to date, no study has simultaneously evaluated the influence of these three biomarkers on MS disability progression. Moreover, most studies investigating brain lesion measures have been conducted using 3T MRI (5,6). Compared to lower field strength, 7T MRI is considered the gold standard for the in vivo detection of CLs (9) and PRLs (10), allowing for the identification of a significantly higher number of lesions (11) and improving the differentiation among different CL types (12,13). Assessing lesion loads with highly sensitive methods may improve the reliability of the association between clinical outcomes and imaging predictors.

Expanding on our previous work (7), this study evaluated, in a heterogenous MS cohort, brain lesion biomarkers from 7T acquisitions and brain and spinal cord atrophy from 3T MRI to identify key predictors of baseline disability and the risk of PIRA over 4 years.

## Materials and Methods

### Standard Protocol Approvals, Registrations, and Patient Consents

The study was approved by the Mass General Brigham Institutional Review Board (#2007P001274) and written informed consent in accordance with the Declaration of Helsinki was obtained from all participants before study enrolment.

### Study population

A total of 129 patients patients meeting MS diagnosis (14) criteria and 41 age-matched healthy controls (HC) were prospectively enrolled in the study between 2010 and 2024. Of the 129 patients, 111 had taken part in two previous studies (7,15). These prior articles used a machine learning approach to identify predictors of EDSS progression and cortical atrophy, whereas in this study we incorporated additional imaging metrics, refined the clinical outcome to focus on PIRA, and employed conventional statistical methods. Inclusion and exclusion criteria are reported in Figure 1. All patients were either on stable disease modifying therapy (DMT) or without treatment for at least three months.

**Figure 1.**
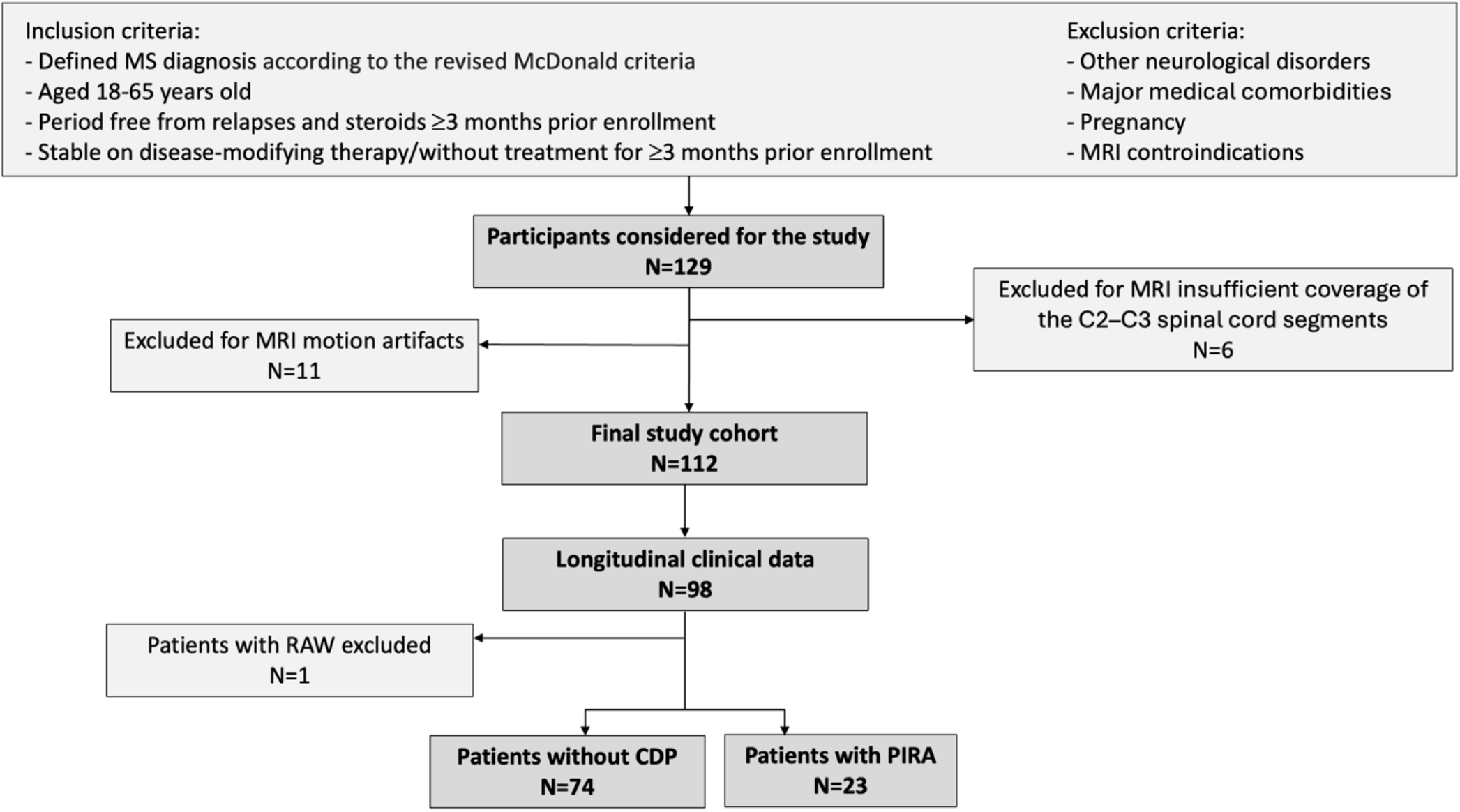
Flowchart detailing the inclusion and exclusion criteria and the selection process of the MS study cohort. CDP = confirmed disability progression; MS = multiple sclerosis; PIRA = progression independent of relapse activity; RAW = relapse-associated worsening.

### Clinical Assessment

All patients underwent a neurological examination within two weeks of the MRI scan (baseline timepoint) with neurological disability assessed using the EDSS (16). Neurological evaluations were regularly peformed every 6-12 months during a mean follow-up period of 4.0 years (SD ± 1.9) in 98/112 patients (75 relapsing-remitting [RRMS], 23 secondary progressive [SPMS]). Confirmed disability progression (CDP) was defined as a disability increase in EDSS score, confirmed at least after 6 months, of: (1) ≥1.5 points if the baseline EDSS was 0, (2) ≥1.0 point if the baseline EDSS was between 1.0 and 5.5, and (3) ≥0.5 points if the baseline EDSS was greater than 5.5. During the follow-up, CDP events were classified as PIRA if no relapses occurred i) between the EDSS increase and the preceding reference visit (conducted at least 90 days earlier), and (2) between the EDSS increase and the confirmation of disability progression. Since the study aimed to identify predictors of PIRA, patients with documented clinical relapses that resulted in disability worsening (RAW) were excluded from the longitudinal analysis. Patients were then classified as either patients who developed PIRA or patients without CDP. The DMT class in which the patient was on was classified as either none, low-moderate efficacy therapy (LMET), or high-efficacy therapy (HET) (17).

### MRI Protocol

All study participants underwent two imaging sessions within a week on a 7-T and a 3-T MRI human scanners (Siemens, Erlangen, Germany) using 32 channel coils to acquire the following brain sequences: (i) 7-T 2D fast low-angle shot (FLASH) T2*-weighted spoiled gradient-echo (GRE) image (repetition time/echo time [TR/TE] 1700/21.8 msec, flip angle = 55°, two slabs of 40 slices each to cover the supratentorial brain, resolution = 0.33 × 0.33 × 1 mm^3^ (25% gap), bandwidth = 335 Hz/pixel, acquisition time for each slab = ∼10 min) yielding magnitude and phase images; (ii) 7-T T1-weighted 3D magnetization-prepared rapid acquisition gradient echo sequence (MPRAGE, TR/TE = 2600/3.26 msec, inversion time [TI] = 1100 msec, flip angle = 9°, field of view = 174 × 192 mm^2^, resolution = 0.60 × 0.60 × 1.5 mm^3^, bandwidth = 200 Hz/pixel, acquisition time = 5.5 min) for coregistration of 7T GRE data with cortical surfaces generated from 3T anatomical acquisitions; (iii) 3-T T1-weighted 3D magnetization-prepared rapid acquisition with multiple gradient echoes (MEMPRAGE) [TR/TI= 2530/1200 msec, TE = (1.7, 3.6, 5.4, 7.3) msec, flip angle = 7°, field of view = 230 × 230 mm^2^, resolution = 0.9 × 0.9 × 0.9 mm^3^, bandwidth = 651 Hz/pixel, acquisition time = ∼6.5 min]), covering the brain and the SC until the segment C4.

### MRI data processing

#### Lesion segmentation

Cortical and white matter (WM) lesions were segmented on magnitude images from 7T T2* scans by consensus of two experienced raters, one radiologist (CAT), and one neurologist (CM), each with over 15 years of experience in neuroimaging analysis, using a semi-automated tool in 3D Slicer version 4.2.0. Cortical lesions were defined as as clearly hyperintense lesions within the cortex, potentially extending towards the WM, covering at least 3 voxels across two consecutive slices, including subpial, intracortical and leukocortical lesion subtypes (13). Paramagnetic rim lesions were defined as WM lesions having a T2-hyperintense core, surrounded by a paramagnetic rim on phase images, along at least 2/3 of the lesion perimeter, and either lacking gadolinium enhancement or present in another MRI scan occurring at least 3 months either before or after the initial scan, according to recommended guidelines (10). Based on this criterion, WM lesions were classified as either non-rim or PRLs. Figure 2 shows examples of the segmentation of different brain lesion phenotypes. Lesion masks from 7T T2* were registered onto the 3T anatomical FreeSurfer reconstructions using a boundary-based registration method as previously detailed (18) (FreeSurfer version 6.0) and lesion count and volumes were quantified using fslstats (FMRIB Software Library, FSL, v. 5.0, Oxford, UK).

**Figure 2.**
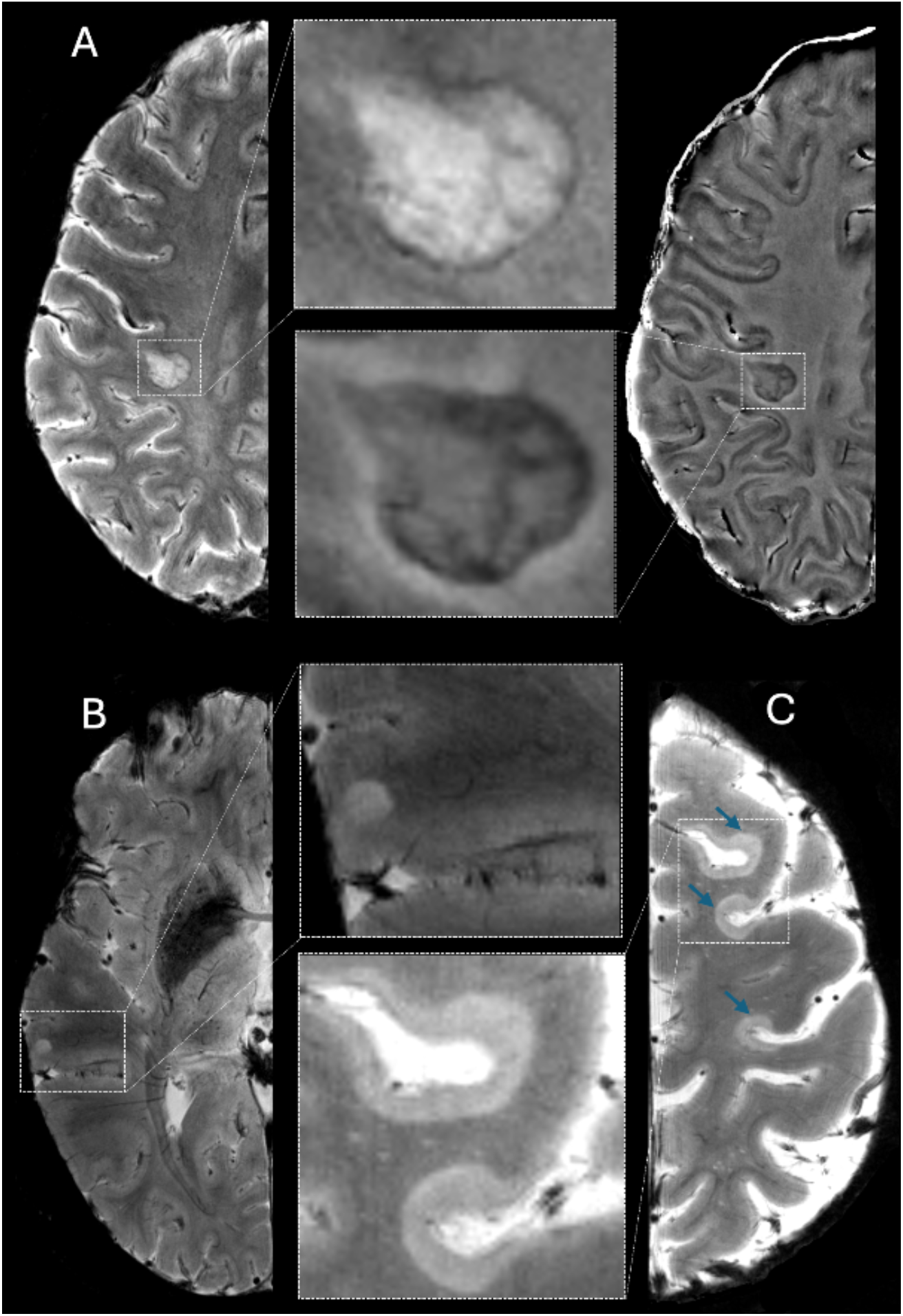
Examples of paramagnetic rim and cortical lesions. **(A)** shows a PRL on 7T T2* magnitude (left) and unwrapped phase (right) images, along with their respective magnifications. Examples of cortical lesions are shown on a 7T T2* magnitude images, namely one leukocortical lesion **(B)** and three type IV subpial lesions **(C, blue arrows)**, also magnified. PRL = paramagnetic rim lesions.

#### Brain volume and spinal cord area measurements

Cortical thickness and brain WM volume were quantified on 3T three-dimensional T1-weighted MPRAGE images, after lesion filling, using FreeSurfer version 6.0. Brain WM volume was normalized by the total intracranial volume. For SC morphological analysis, we measured the mean cross-sectional area (CSA) across C2-C3 vertebral levels through the DeepSeg algorithm from the Spinal Cord Toolbox (SCT) version 6.1 (19) using 3T T1-weighted images as input. The C2-C3 intervertebral disk was manually labeled in each image to ensure optimal placement, and all pipeline steps were manually reviewed. Figure 3 shows SC C2-C3 segmentation and CSA estimation.

**Figure 3.**
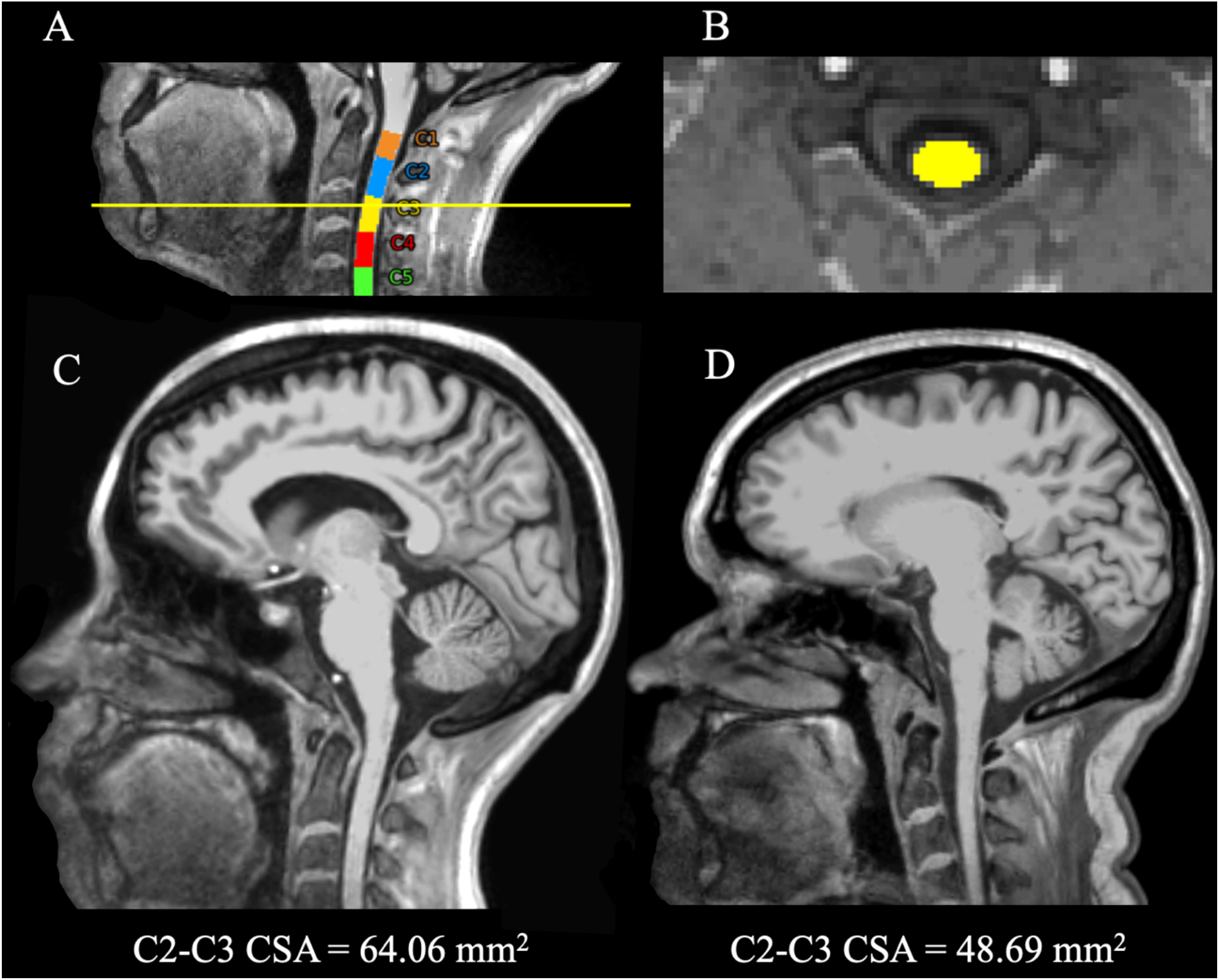
Example of automatic segmentation of the cervical spinal cord on 3T T1-weighted sagittal **(A)** and axial **(B)** planes to estimate the C2-C3 CSA, using Spinal Cord Toolbox version 6.1. Sagittal T1-weighted images show the difference in C2-C3 CSA between a HC **(C)** and a SPMS patient (disease duration = 15 years) **(D)**. CSA = cross-sectional area; HC = healthy control; SPMS = secondary progressive multiple sclerosis.

### Statistical analysis

Statistical analyses were performed using IBM SPSS Statistics (Version 25.0). Differences between groups (i.e. MS patients vs HC; RRMS vs SPMS patients; patients without CDP vs patients with PIRA) were analyzed using the chi-squared test for categorical variables, the independent-samples t-test for parametric continuous variables, and the Mann–Whitney test for nonparametric continuous variables. Since the occurrence of PIRA depends on the length of the follow-up interval, longitudinal analyses were thoroughly adjusted for follow-up time. We investigated the association of imaging metrics with baseline EDSS using multivariable linear regressions adjusted for age, sex, and DMT class, and with PIRA using multivariable logistic regressions adjusted for age, sex, DMT class, and clinical follow-up interval. To determine which baseline features were independently associated with the outcome when considered together, we performed a multivariable stepwise logistic regression to predict PIRA (vs. non-CDP). Collinearity between predictors in regression models was excluded based on the Variance Inflation Factor (VIF). A receiver operating characteristic (ROC) curve was used to determine the optimal cut-off volume of the predictive biomarker that best distinguishes patients at risk of PIRA from those who are not. A p-value of 0.05 was considered statistically significant.

## Results

### Demographic, clinical, and MRI characteristics of the study population

Out of 129 MS patients, data from 11 participants were excluded due to motion artifacts, and 6 due to insufficient coverage of the C2–C3 spinal cord segments within the brain MRI field of view. A total of 112 patients were included in the final analysis. At baseline, 72 (64%) had at least one PRL, and 107 (95%) had at least one CL.

After a mean follow-up period of 4.0 (SD 1.9) years, 9 out of 98 patients (all with RRMS) experienced at least one relapse during the follow-up; however, only one of them had RAW and was excluded from the longitudinal analysis. The final longitudinal cohort included 97 patients (74 RRMS, 23 SPMS), of whom 23 (24%) with PIRA (13 RRMS, 10 SPMS). Table 1 summarizes the main demographic, clinical, and MRI features of HC and MS patients, categorized by clinical phenotype and longitudinal disability trajectory.

**Table 1.**
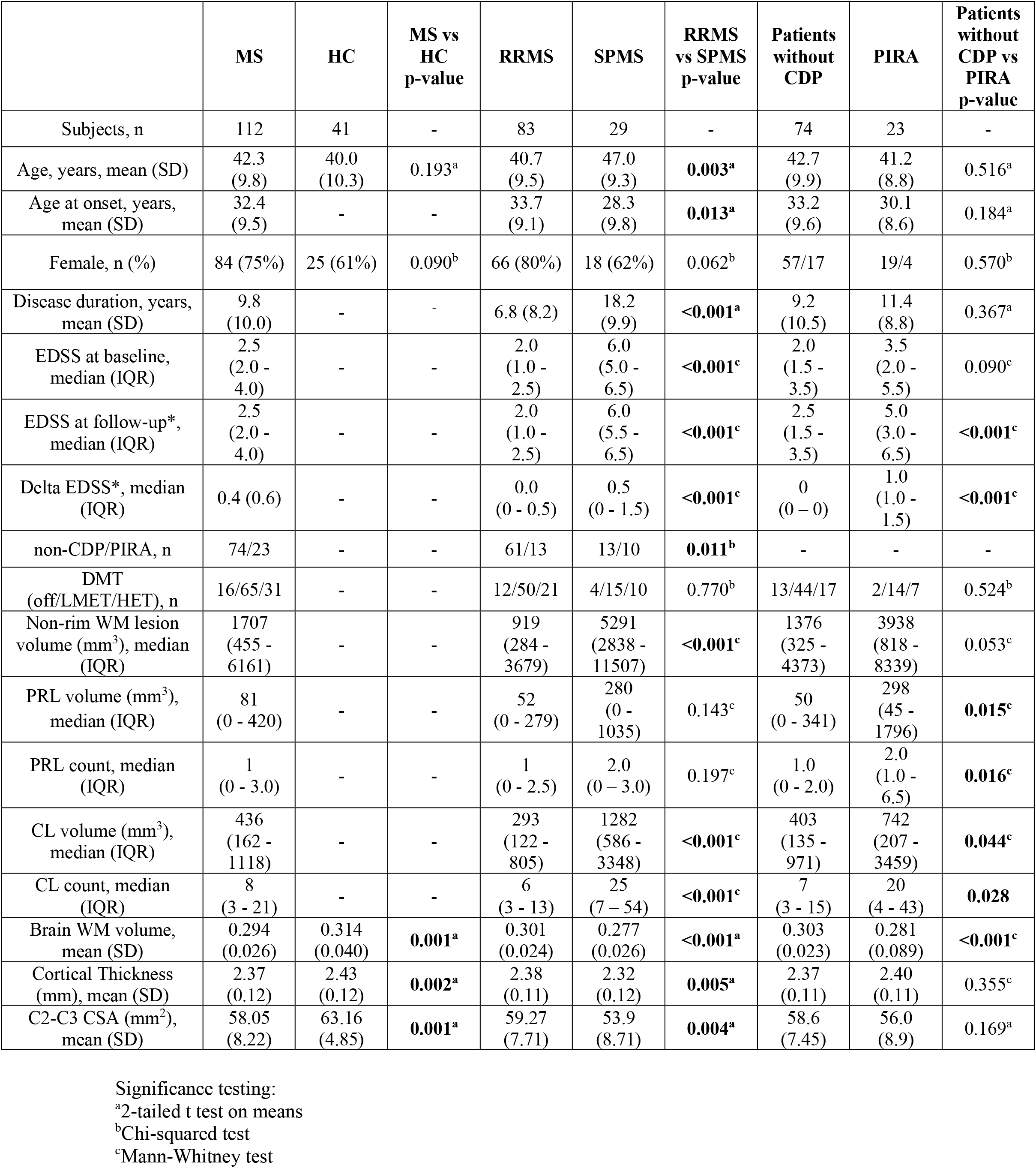
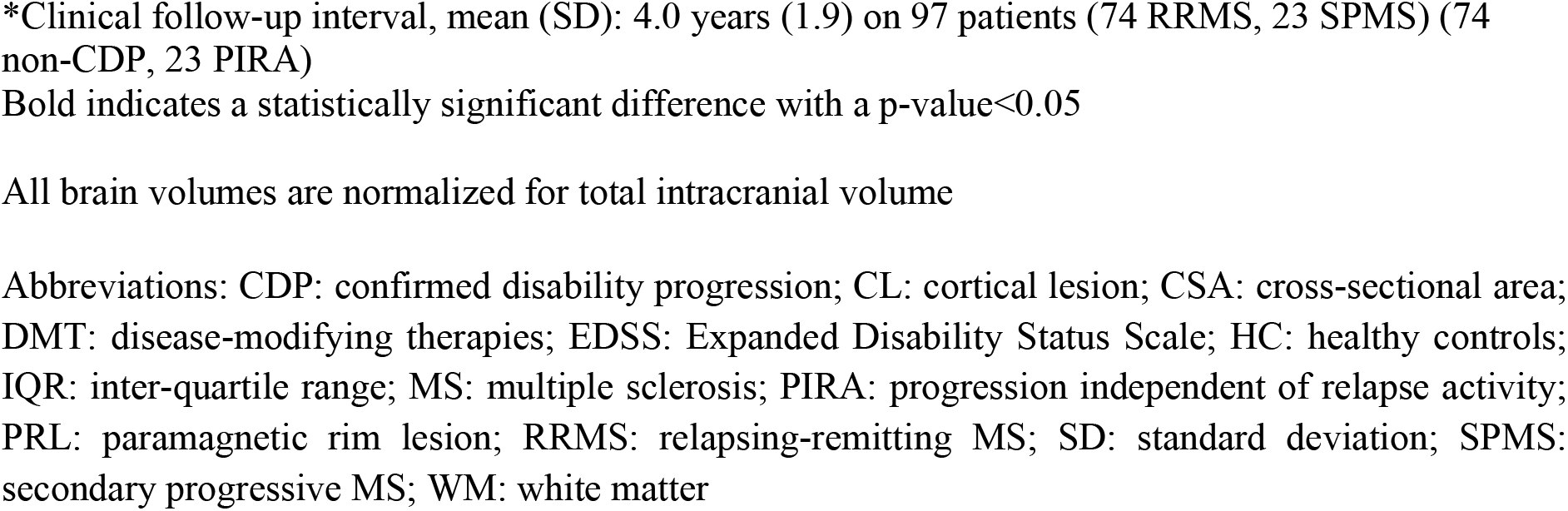
Demographics, clinical, and radiological characteristics.

|  | MS | HC | MS vs<br>HC<br>p-value | RRMS | SPMS | RRMS<br>vs SPMS<br>p-value | Patients<br>without<br>CDP | PIRA | Patients<br>without<br>CDP vs<br>PIRA<br>p-value |
| --- | --- | --- | --- | --- | --- | --- | --- | --- | --- |
| Subjects, n | 112 | 41 | - | 83 | 29 | - | 74 | 23 | - |
| Age, years, mean (SD) | 42.3<br>(9.8) | 40.0<br>(10.3) | 0.193<sup>a</sup> | 40.7<br>(9.5) | 47.0<br>(9.3) | **0.003<sup>a</sup>** | 42.7<br>(9.9) | 41.2<br>(8.8) | 0.516<sup>a</sup> |
| Age at onset, years,<br>mean (SD) | 32.4<br>(9.5) | - | - | 33.7<br>(9.1) | 28.3<br>(9.8) | **0.013<sup>a</sup>** | 33.2<br>(9.6) | 30.1<br>(8.6) | 0.184<sup>a</sup> |
| Female, n (%) | 84 (75%) | 25 (61%) | 0.090<sup>b</sup> | 66 (80%) | 18 (62%) | 0.062<sup>b</sup> | 57/17 | 19/4 | 0.570<sup>b</sup> |
| Disease duration, years,<br>mean (SD) | 9.8<br>(10.0) | - | - | 6.8 (8.2) | 18.2<br>(9.9) | **<0.001<sup>a</sup>** | 9.2<br>(10.5) | 11.4<br>(8.8) | 0.367<sup>a</sup> |
| EDSS at baseline,<br>median (IQR) | 2.5<br>(2.0 -<br>4.0) | - | - | 2.0<br>(1.0 -<br>2.5) | 6.0<br>(5.0 -<br>6.5) | **<0.001<sup>c</sup>** | 2.0<br>(1.5 -<br>3.5) | 3.5<br>(2.0 -<br>5.5) | 0.090<sup>c</sup> |
| EDSS at follow-up*,<br>median (IQR) | 2.5<br>(2.0 -<br>4.0) | - | - | 2.0<br>(1.0 -<br>2.5) | 6.0<br>(5.5 -<br>6.5) | **<0.001<sup>c</sup>** | 2.5<br>(1.5 -<br>3.5) | 5.0<br>(3.0 -<br>6.5) | **<0.001<sup>c</sup>** |
| Delta EDSS*, median<br>(IQR) | 0.4 (0.6) | - | - | 0.0<br>(0 - 0.5) | 0.5<br>(0 - 1.5) | **<0.001<sup>c</sup>** | 0<br>(0 - 0) | 1.0<br>(1.0 -<br>1.5) | **<0.001<sup>c</sup>** |
| non-CDP/PIRA, n | 74/23 | - | - | 61/13 | 13/10 | **0.011<sup>b</sup>** | - | - | - |
| DMT<br>(off/LMET/HET), n | 16/65/31 | - | - | 12/50/21 | 4/15/10 | 0.770<sup>b</sup> | 13/44/17 | 2/14/7 | 0.524<sup>b</sup> |
| Non-rim WM lesion<br>volume (mm<sup>3</sup>), median<br>(IQR) | 1707<br>(455 -<br>6161) | - | - | 919<br>(284 -<br>3679) | 5291<br>(2838 -<br>11507) | **<0.001<sup>c</sup>** | 1376<br>(325 -<br>4373) | 3938<br>(818 -<br>8339) | 0.053<sup>c</sup> |
| PRL volume (mm<sup>3</sup>),<br>median (IQR) | 81<br>(0 - 420) | - | - | 52<br>(0 - 279) | 280<br>(0 -<br>1035) | 0.143<sup>c</sup> | 50<br>(0 - 341) | 298<br>(45 -<br>1796) | **0.015<sup>c</sup>** |
| PRL count, median<br>(IQR) | 1<br>(0 - 3.0) | - | - | 1<br>(0 - 2.5) | 2.0<br>(0 - 3.0) | 0.197<sup>c</sup> | 1.0<br>(0 - 2.0) | 2.0<br>(1.0 -<br>6.5) | **0.016<sup>c</sup>** |
| CL volume (mm<sup>3</sup>),<br>median (IQR) | 436<br>(162 -<br>1118) | - | - | 293<br>(122 -<br>805) | 1282<br>(586 -<br>3348) | **<0.001<sup>c</sup>** | 403<br>(135 -<br>971) | 742<br>(207 -<br>3459) | **0.044<sup>c</sup>** |
| CL count, median<br>(IQR) | 8<br>(3 - 21) | - | - | 6<br>(3 - 13) | 25<br>(7 - 54) | **<0.001<sup>c</sup>** | 7<br>(3 - 15) | 20<br>(4 - 43) | **0.028** |
| Brain WM volume,<br>mean (SD) | 0.294<br>(0.026) | 0.314<br>(0.040) | **0.001<sup>a</sup>** | 0.301<br>(0.024) | 0.277<br>(0.026) | **<0.001<sup>a</sup>** | 0.303<br>(0.023) | 0.281<br>(0.089) | **<0.001<sup>c</sup>** |
| Cortical Thickness<br>(mm), mean (SD) | 2.37<br>(0.12) | 2.43<br>(0.12) | **0.002<sup>a</sup>** | 2.38<br>(0.11) | 2.32<br>(0.12) | **0.005<sup>a</sup>** | 2.37<br>(0.11) | 2.40<br>(0.11) | 0.355<sup>c</sup> |
| C2-C3 CSA (mm<sup>2</sup>),<br>mean (SD) | 58.05<br>(8.22) | 63.16<br>(4.85) | **0.001<sup>a</sup>** | 59.27<br>(7.71) | 53.9<br>(8.71) | **0.004<sup>a</sup>** | 58.6<br>(7.45) | 56.0<br>(8.9) | 0.169<sup>a</sup> |

Significance testing:
<sup>a</sup>2-tailed t test on means<sup>b</sup>Chi-squared test<sup>c</sup>Mann-Whitney test
\*Clinical follow-up interval, mean (SD): 4.0 years (1.9) on 97 patients (74 RRMS, 23 SPMS) (74 non-CDP, 23 PIRA)
Bold indicates a statistically significant difference with a p-value<0.05
All brain volumes are normalized for total intracranial volume
Abbreviations: CDP: confirmed disability progression; CL: cortical lesion; CSA: cross-sectional area; DMT: disease-modifying therapies; EDSS: Expanded Disability Status Scale; HC: healthy controls; IQR: inter-quartile range; MS: multiple sclerosis; PIRA: progression independent of relapse activity; PRL: paramagnetic rim lesion; RRMS: relapsing-remitting MS; SD: standard deviation; SPMS: secondary progressive MS; WM: white matter

Compared to HC, patients with MS had lower normalized brain WM volume (p = 0.001), cortical thinning (p = 0.002), and reduced SC C2-C2 CSA (p = 0.001). Compared to RRMS, SPMS patients were older (p = 0.003), had longer disease duration (p < 0.001), and more severe EDSS scores (p < 0.001). SPMS patients showed worse MRI parameters in all lesion and volumetric metrics (p < 0.005 for all comparisons), except for PRL volume and count.

Compared to patients without CDP, patients with PIRA had significantly higher EDSS at follow-up (p < 0.001), higher PRL volume (p = 0.015) and count (p = 0.016), higher CL volume (p = 0.044) and count (p = 0.028), and lower brain WM volume (p < 0.001).

### MRI biomarkers associated with current MS disability

At baseline, linear regression models (Table 2), adjusted for age, sex, and DMT class, showed significant associations between current EDSS and volumes of non-rimWM lesions (β = 1.2×0^-4^, p < 0.001), CLs (β = 2.9×10^−4^, p = 0.001), brain WM (β = -15.68, p = 0.017) as well as with SC C2-C3 CSA (β = -0.68, p = 0.003). Among covariates, age was the only one significantly associated with baseline EDSS (β =0.069, p = 0.001).

**Table 2.** Multivariable regression analyses of MRI biomarkers with baseline EDSS and PIRA

|  | Baseline EDSS<sup>a</sup> |  |  | PIRA<sup>b</sup> |  |  |  |
| --- | --- | --- | --- | --- | --- | --- | --- |
| MRI variables | R<sup>2</sup> | Unstandardized $\beta$ (95% IC) | P-value | AUC | OR (95% IC) | $\beta$ | P-value |
| Non-rim WM lesion volume | 0.247 | 1.2x10<sup>-4</sup> (0.6 x10<sup>-4</sup> – 1.8 x10<sup>-4</sup>) | **<0.001** | 0.631 | 1.0001 (0.9999 – 1.0002) | 1.0 x10<sup>-4</sup> | 0.052 |
| PRL volume | 0.181 | 1.2x10<sup>-4</sup> (0.3 x10<sup>-4</sup> – 2.7 x10<sup>-4</sup>) | 0.113 | 0.658 | 1.0006 (1.0001 – 1.0010) | 5.7 x10<sup>-4</sup> | **0.030** |
| CL volume | 0.238 | 2.9x10<sup>-4</sup> (1.2 x10<sup>-4</sup> – 4.6 x10<sup>-4</sup>) | **0.001** | 0.633 | 1.0005 (1.0001 – 1.0009) | 5.3 x10<sup>-4</sup> | **0.011** |
| Brain WM volume | 0.198 | -15.68 (-28.61 – -2.76) | **0.017** | 0.238 | 0.97x10<sup>-20</sup> (0.10x10<sup>-30</sup> – 0.95x10<sup>-9</sup>) | -46.07 | **<0.001** |
| Cortical Thickness | 0.179 | -2.72 (-5.91 – 0.46) | 0.094 | 0.569 | 0.991 (0.121 – 1.825) | -2.86 | 0.260 |
| C2-C3 CSA | 0.241 | -0.68 (-0.11 – -0.02) | **0.003** | 0.426 | 0.946 (0.883 – 1.013) | -0.056 | 0.113 |
| age | - | 0.069 (0.030 – 0.107) | **0.001** | - | 0.979 (0.939 – 1.031) | -0.21 | 0.428 |
| sex | - | -0.077 (-0.928 – 0.773) | 0.859 | - | 0.608 (0.164 – 2.259) | -0.497 | 0.458 |
| DMT class | - | -0.703 (-1.548 – 0.142) | 0.103 | - | 1.437 (0.622 – 3.323) | 0.343 | 0.397 |
| follow-up duration | - | - | - | - | 1.416 (1.060 – 1.892) | 0.348 | **0.019** |

<sup>a</sup>Multivariable linear regressions including the single MRI biomarker along with age, sex, and DMT class as independent variables
<sup>b</sup>Multivariable logistic regressions including the single MRI biomarker along with age, sex, DMT class, and follow-up duration as independent variables
Abbreviations: CL: cortical lesion; CSA: cross-sectional area; DMT: disease-modifying therapies; EDSS: Expanded Disability Status Scale; OR = odds ratio; PIRA: Progression independent of relapse activity; PRL: paramagnetic rim lesion; WM: white matter

### MRI predictors of PIRA

Logistic regression models (Table 2), adjusted for age, sex, DMT class, and clinical follow-up time, identified associations between PIRA and PRL volume (odds ratio [OR] = 1.0006, p = 0.030), CL volume (OR = 1.0005, 0.011), and normalized brain WM volume (OR = 0.97×10^−20^, p < 0.001). Among covariates, follow-up duration was the only factor associated with PIRA (OR = 1.416, p = 0.019).

To further explore independent predictors of PIRA, a stepwise forward logistic regression analysis was performed, including age, sex, DMT class, follow-up duration, disease duration, clinical phenotype (RRMS/SPMS), non-rim WM lesion volume, PRL volume, CL volume, WM volume, cortical thickness, and C2–C3 CSA as candidate predictors (all with VIF < 3 to rule out multicollinearity). The CL volume was the only predictor of PIRA retained in the first step of the model (Nagelkerke R^2^ = 0.200, OR = 1.0006 [95% CI: 1.0002–1.0010], β = 0.006, p = 0.005). In the second step, brain WM volume was added, significantly improving model fit (Nagelkerke R^2^ = 0.298, OR = 0.89 × 10−^15^ [95% CI: 0.43 × 10−^27^ – 0.002], β = –34.6, p = 0.017). Despite this, CL volume remained an independent contributor (p = 0.038). Based on the coordinates of the ROC curve (Fig. 4), patients with a baseline CL load > 403 mm^3^ progressed in half of the cases (70% sensitivity, 50% specificity) within a mean follow-up of 4 years.

**Figure 4.**
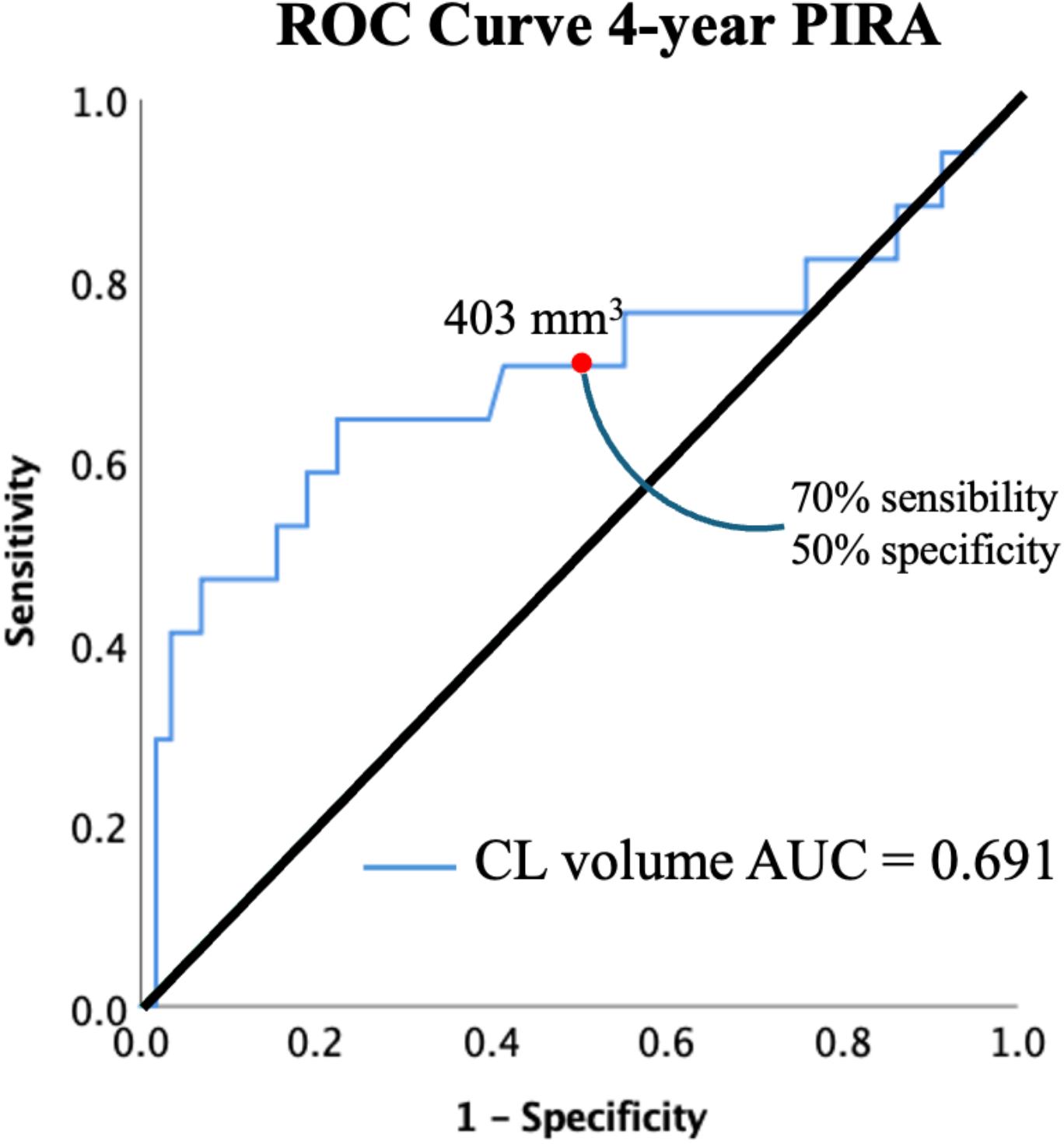
ROC curve to quantify the CL load required to be exposed to the risk of PIRA. It revealed that 50% of the patients with a baseline CL volume of at least 403 mm^3^ progressed in EDSS within a mean period of 4 years, with 70% sensitivity. AUC = area under the curve; CL = cortical lesion; EDSS = expanded disability status scale; PIRA = progression independent of relapse activity; ROC = receiver operating characteristic.

## Discussion

In this 7T and 3T MRI study in a large cohort of individuals with MS, we evaluated the contribution of SC and brain imaging biomarkers to baseline clinical disability and PIRA. We found that cervical SC atrophy is a major indicator of the present EDSS score, but plays a limited role in predicting PIRA compared to brain biomarkers. The contribution of WM lesions depends on the presence of smoldering activity, with PRLs providing information on the future disability trajectory and non-rim WM lesions reflecting baseline EDSS. Brain WM atrophy emerged as index of current disability and risk of PIRA. Finally, we found that cortical lesion load, as assessed by 7T MRI, is the strongest driver of PIRA over a 4-year period, as well as showing an association with the current EDSS.

Spinal cord degeneration is widely recognized as a valid predictor of physical disability and disease progression in MS (20,21). A recent study identified baseline C2–C3 CSA as an independent predictor of time to PIRA in a cohort of 445 MS patients clinically followed over 4 years (5). In contrast, although patients who experienced PIRA in our cohort tended to have lower mean cervical SC CSA values compared to clinically stable individuals, baseline C2–C3 CSA did not significantly predict PIRA, possibily due to limited statistical power from the smaller sample size. Nonetheless, our integrated analysis of brain and spinal cord measures revealed strong associations between PIRA and brain biomarkers, suggesting a predominant role of brain pathology, rather than SC atrophy, in its prediction.

Brain non-rim WM lesions represent the majority of the MS lesions, although disability can progress regardless of their accumulation (22). This suggests that other types of tissue damage may be key drivers of disease progression. Accordingly, we found that baseline non-rim WM lesion volume was primarily associated with current EDSS scores but did not demonstrate a significant predictive value for future disability. In contrast, PRLs represent a manifestation of smoldering MS, characterized by persistent inflammation at the borders of MS plaques with a rim of iron-rich active microglia/macrophages, and have been associated with a more severe disease course and disability accumulation (23,10). Indeed, our data revealed that baseline PRL volume is a robust predictor of the 4-year PIRA. Being associated with disability progression but not with baseline EDSS, PRLs clearly differentiate their prognostic significance from that of non-rim WM lesions. It is conceivable that PRLs impact disability accumulation as long as their instrinsic smoldering activity persists.

Longitudinal quantitative susceptibility mapping (QSM) studies have shown that QSM value in the chronic active rim gradually decreases after the fourth year of their formation (24), and PRLs might eventually convert into non-rim WM lesions, as part of a continuum. At that point, their prognostic contribution might remain limited to the overall EDSS score, but not necessarily to PIRA.

Interestingly, we found that PRL and CL volumes share an association with disability progression, with the latter having the highest impact on PIRA. There is evidence that CL volume increases significantly over time in individuals with ≥3 PRLs (25), who might represent patients with a more aggressive disease progression and both cortical and WM inflammatory profiles. Cortical MS lesions are strongly associated with disability progression, and in some studies, they are even more predictive than WM lesions, particularly when assessed using ultra-high field MRI (7,25,26). While the detection of leukocortical lesions at 3T MRI is comparable to that at 7T, the contribution of ultra-high field imaging to the identification of intracortical and subpial lesions is substantial, revealing up to ten times more lesions than 3T images (27). Histopathological studies have revealed that meningeal inflammation, particularly in the form of lymphoid-like structures with B-cell clonal expansion and Epstein-Barr virus (EBV)-reactivated B cells, is strongly associated with cortical pathology (28,29). These ectopic lymphoid follicles are predominantly observed in progressive MS and have been linked to widespread subpial grey matter demyelination. Thus, the link between CLs and PIRA may lie in compartmentalized meningeal inflammation that sustains cortical demyelination. The detection of CLs may therefore provide indirect but clinically meaningful insights into both cortical and meningeal immune activity in MS.

Our study also confirmed that brain WM atrophy is highly associated with current disability and PIRA (7,30). In contrast, cervical SC CSA was not retained as significant predictor of PIRA. Brain and SC volumetry has become a valuable tool for understanding MS progression, reflecting neuroaxonal loss occurred in the CNS. This may represent the effect of subtle chronic inflammation that persists throughout the disease course, leading to greater atrophy in patients with more pronounced smoldering activity. In particular, brain WM may be more closely associated with compartmentalized inflammation, given its proximity to key sources of such inflammation, including enlarged choroid plexuses (31), PRLs (32), and diffuse microglial activation within the normal-appearing WM (33).

However, the present study builds upon our previous machine learning work (7), which identified PRLs, leukocortical lesion volumes, and brain WM volume as principal predictors of EDSS progression(7). In particular, we: (i) incorporated SC atrophy as an additional, albeit minor contributor to clinical progression; (ii) refined the clinical outcome by isolating PIRA, thus capturing the clinical manifestation of smoldering MS; and (iii) validated our findings using conventional statistical approaches, thereby reinforcing the robustness of the observed associations.

This work has some limitations. We did not assess the contribution of SC lesions to neurologic disability. However, a previous study with 3-year clinical follow-up did not show any correlation between baseline SC lesion number and change in disability measures (25). Another limitation is that clinical phenotype and disease duration, despite their known relevance to MS disability, were not included as covariates in the multivariable models. When tested, they strongly predicted EDSS and PIRA, limiting our ability to assess the independent predictive value of imaging markers. However, they were included in the stepwise logistic regression model, but neither was retained in the models. Finally, follow-up duration varied across patients in our cohort, with a mean of 4 years. Although follow-up time was included as a covariate, this heterogeneity may have influenced the detection of PIRA events, which are inherently time-dependent.

In conclusion, our 7T and 3T MRI study maps the clinical trajectory of numerous SC and brain biomarkers of disability in patients with MS. The CL load has proven to be the strongest predictor of PIRA within 4 years, underscoring the necessity for new imaging techniques able to increase the sensibility for CLs in the clinical setting.

## Data Availability

All data produced in the present study are available upon reasonable request to the authors

## Study Funding

This work was supported by grants from the National Multiple Sclerosis Society (NMSS 4281-RG-A1, NMSS RG 4729A2/1 and NMSS RG 1802-30468), National Institutes of Health R01NS078322-01-A1, United States Army W81XWH-13-1-0122, and FISM 22/R-Multi/028

## Disclosure

Miscioscia A: nothing to disclose. Treaba CA has received research support from Genentech. Barbuti E: nothing to disclose. Barletta V: nothing to disclose. Sloane J has consulted for Novartis, Biogen, Cellgene, Genentech. Klawiter E has received consulting fees from EMD Serono, Genentech, INmune Bio, Myrobalan Therapeutics, OM1and TG Therapeutics, and received research funds from Abbvie, Biogen, and Genentech. Cohen-Adad J: nothing to disclose. Gallo P: nothing to disclose. Pantano P: nothing to disclose. Mainero C has received research support from Genentech.

## Acknowledgment

We would like to thank the patients and their relatives for their participation and support, as well as the hospital staff for their assistance in making this study possible.

